# Machine Learning for Pattern Detection in Cochlear Implant FDA Adverse Event Reports

**DOI:** 10.1101/2020.04.30.20086660

**Authors:** Matthew G. Crowson, Amr Hamour, Vincent Lin, Joseph M. Chen, Timothy C. Y. Chan

## Abstract

**Importance:** The United States Food & Drug Administration (FDA) passively monitors medical device performance and safety through submitted medical device reports (MDRs) in the Manufacturer and User Facility Device Experience (MAUDE) database. These databases can be analyzed for patterns and novel opportunities for improving patient safety and/or device design.

**Objectives:** The objective of this analysis was to use supervised machine learning to explore patterns in reported adverse events involving cochlear implants.

**Design:** The MDRs for the top three CI manufacturers by volume from January 1^st^ 2009 to August 30^th^ 2019 were retained for the analysis. Natural language processing was used to measure the importance of specific words. Four supervised machine learning algorithms were used to predict which adverse event narrative description pattern corresponded with a specific cochlear implant manufacturer and adverse event type - injury, malfunction, or death.

**Setting:** U.S. government public database.

**Participants:** Adult and pediatric cochlear patients.

**Exposure:** Surgical placement of a cochlear implant.

**Main Outcome Measure:** Machine learning model classification prediction accuracy (% correct predictions).

**Results:** 27,511 adverse events related to cochlear implant devices were submitted to the MAUDE database during the study period. Most adverse events involved patient injury (n = 16,736), followed by device malfunction (n = 10,760), and death (n = 16). Submissions to the database were dominated by Cochlear Corporation (n = 13,897), followed by MedEL (n = 7,125), and Advanced Bionics (n = 6,489). The random forest, linear SVC, naïve Bayes and logistic algorithms were able to predict the specific CI manufacturer based on the adverse event narrative with an average accuracy of 74.8%, 86.0%, 88.5% and 88.6%, respectively.

**Conclusions & Relevance:** Using supervised machine learning algorithms, our classification models were able to predict the CI manufacturer and event type with high accuracy based on patterns in adverse event text descriptions.

**Level of evidence:** 3

## INTRODUCTION

The United States Food & Drug Administration (FDA) passively monitors medical device performance and safety through submitted medical device reports (MDRs) in the Manufacturer and User Facility Device Experience (MAUDE) database.^1,2^ The MDRs contain the type of adverse event (i.e., injury, device malfunction or death), the device, manufacturer and a free text narrative of the event. A theoretical advantage of using a centralized device database over a single-institution’s experience is a wider breadth and depth of incidents including all manufacturers of approved medical devices in all fields of medicine and surgery. Frequently cited analyses of cardiovascular devices (e.g., catheters and stents), obstetrical devices, and robots have produced insights into the nature and frequency of device-related adverse events using the MAUDE database.^3–9^

Machine learning algorithms are a subdomain of artificial intelligence and are used to ‘learn’ patterns in large, heterogeneous data to generate predictions. These predictions can augment human decision making and can uncover novel relationships in large datasets that are not observable using traditional statistical techniques. Natural language processing (NLP) - a subset of artificial intelligence - is concerned with the autonomous deduction of meaning and intent from human language using algorithms. Freeform narrative text is ubiquitous in clinical and research settings, and significant recent interest has been established in applying NLP coupled with machine learning to unlock embedded patterns and insights. Successful efforts using NLP with narrative clinical data have been applied to structure narratives within electronic health records,^10–20^ mining drug adverse effect incidence from social media platforms,^21^ pharmaceutical and vaccine safety surveillance,^22–30^ automatic identification of complications during procedures,^31–33^ and patient safety event detection.^34–39^

The MAUDE MDR database has been analyzed to assess cochlear implant (CI) electrode migration,^40^ and temporal trends in CI device failure, surgical site infections, and other adverse events related to CI.^41^ However, as of the writing of this manuscript, no prior analyses used machine learning and natural language processing techniques to analyze the narrative text in CI adverse event reports. Considering the different methodologic approach inherent to machine learning, we hypothesized that we may be able provide additional insights in the study of CI adverse event reports. Specifically, the primary objective of this analysis was to use supervised machine learning to predict the manufacturer of a CI device based on the adverse event narrative text. A secondary objective was to explore patterns in the narrative text by the type of adverse event. By examining patterns in adverse events related to specific manufacturers and types of adverse events, we hoped to mine for novel opportunities for device improvement and patient safety.

## METHODS

This study involved publicly available data, so a review through the Sunnybrook Health Sciences Center institutional review board (IRB) was not undertaken.

### Manufacturer and User Facility Device Experience (MAUDE) Database

e mined the MAUDE database for all available CI-related MDRs submitted to the FDA over the period from January 1st 2009 to August 30th 2019. The MAUDE database contains event type, date, device name, event narrative description and manufacturer in each MDR. The narrative description in the MDRs for the top three CI manufacturers by volume of adverse events were retained for the analysis.

### Data Processing

Free-text narrative data needs to be processed before it can be used as input for a machine learning algorithm in natural language processing tasks. The narrative text from individual adverse event reports was cleaned by removing blank rows, standardizing text case, removing common ‘stop-words’ (e.g., “a,” *“and,” “but,” “how,” “or,”* “*the*,” “who,” and *“what’)* and non-numeric characters and punctuation. We lemmatized individual words to remove inflectional endings and retain the ‘base’ dictionary form of the word. After the text data were cleaned, we used term frequency-inverse document frequency (TF-IDF) natural language processing to measure the importance of specific words in each adverse event proportional to its frequency within the entry and the entire database. The importance value for words are converted into a specific numerical vector values (i.e., vectorization) that allow the comparison between specific words.

### Machine Learning Algorithms

After the text data were prepared, we deployed a five-fold cross validation. Cross-validation is a resampling procedure used to evaluate machine learning models. In five-fold cross validation, the dataset is randomly sampled to generate five different folds whereby each fold is used a testing fold during an iterative training-testing process. In each fold, we used four subsets (80% of the total dataset) to train the model and the remaining subset (20% of the total dataset) to test model performance on data unseen by the model during training. This process was repeated four additional times, and the final accuracy was the aggregate accuracy over all five of the test sets.

The analytic goal of the machine learning algorithms was to classify the narrative text from individual adverse events as i) belonging to one of the three manufacturers, and ii) corresponding to the type of adverse event report – injury, malfunction, or death – in separate analyses (**Figure 1**). Based on their successful use in other applications of natural language processing text classification, we deployed a random forest classifier (200 estimators with a maximum depth of 3), linear support vector classifier (SVC), multinomial Naïve Bayes (mNB) classifier, and multinomial logistic regression classifier (Python *scikit-learn 0.21.3* package).^42 43^ Several metrics were calculated to evaluate the individual classification performance for each model including precision, recall, and the F_1_ score. The precision is the ratio of true positives (TP) over the sum of the true positives (TP) and false positives (FP): TP/(TP+FP). The model recall is the ratio of TP over the sum of TP and false negatives (FN): TP/(TP+FN). The F_1_ score is the weighted average of the precision and recall: 2 x (precision x recall) / (precision + recall).

**Figure 1.**
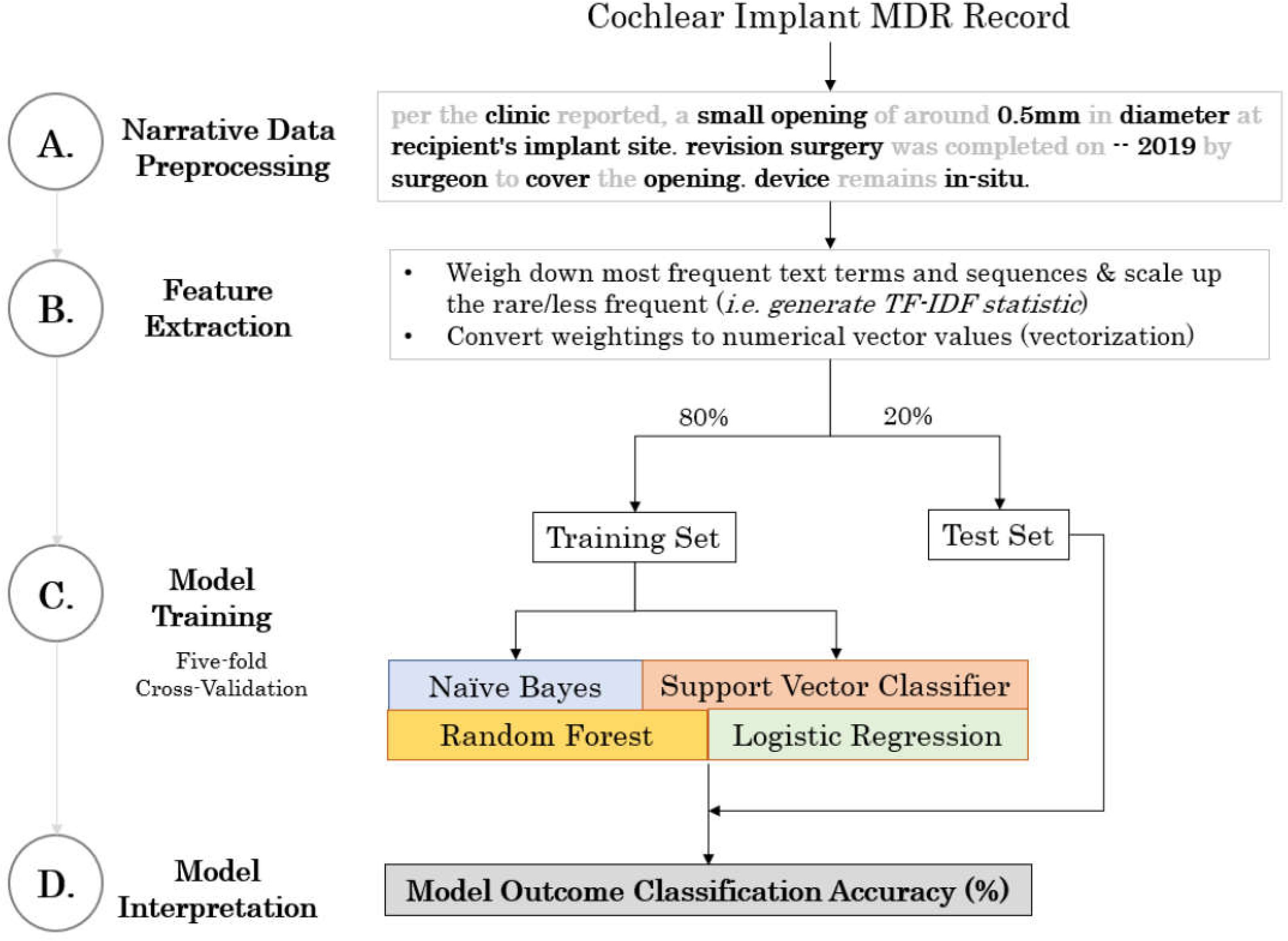
Analytic workflow for preparing and analyzing MAUDE-derived cochlear implant medical device reports.

### Model Interpretation

To illustrate the utility of the machine learning algorithms in performing classification of adverse event narrative text, we asked the best performing algorithm to predict the manufacturer from novel narrative text generated by the authors as examples of common device-related adverse events. The models were interpreted using the interpretable model-agnostic explanations (LIME) package *(version 0.1.1.36)^44^* The LIME approach provides a qualitative understanding between the input variables and the output generated by the classifier algorithm. We deployed LIME to provide a probability (%) of the text sample belonging to a given manufacturer along with the most important text features that account for each prediction. The ELI5 package *(version 0.9.0)* was used to compute the weights of the most common positive and negative predictors for each manufacturer. The numerical weights represent how much each word contributed to the final manufacturer category prediction.

### Hardware & Software Environment

The model training and analyses were completed on a custom-built desktop computer with an AMD Ryzen Threadripper 1950X processor, 32GB of read-only memory (RAM), and a GeForce RTX 2070 8GB graphics card. Data processing and algorithm computation was completed using the Python version 3.6 environment running in an Ubuntu Linux operating system.

## RESULTS

27,512 cochlear implant adverse events from the top three most common manufacturers were submitted to the MAUDE database from January 1st, 2009 to August 30th, 2019. Submissions to the database were led by Cochlear Corporation (n = 13,897; 50.5%), followed by Med-EL (n = 7,125; 25.9%), and Advanced Bionics (n = 6,489; 23.6%). Most adverse events involved patient injury (n = 16,736; 60.8%), followed by device malfunction (n = 10,760; 39.1%), and death (n = 16; 0.1%).

### Machine learning algorithm classification accuracy performance

Our final adverse event report narrative database contained 1,275,668 words with a vocabulary size of 7,858 unique words, and a maximum adverse event report narrative length of 477 words. Based on the free text narrative data alone, the random forest, linear SVC, naïve Bayes and logistic algorithms were able to predict the specific CI manufacturer based on the adverse event narrative with an average accuracy of 74.8%, 86.0%, 88.5% and 88.6%, respectively, after five-fold cross validation (**Figure 2**). For the adverse event type classification task, the random forest, linear SVC, naïve Bayes and logistic algorithms produced an average accuracy of 69.7%, 84.0%, 85.4% and 85.5%, respectively (**Figure 3**).

**Figure 2.**
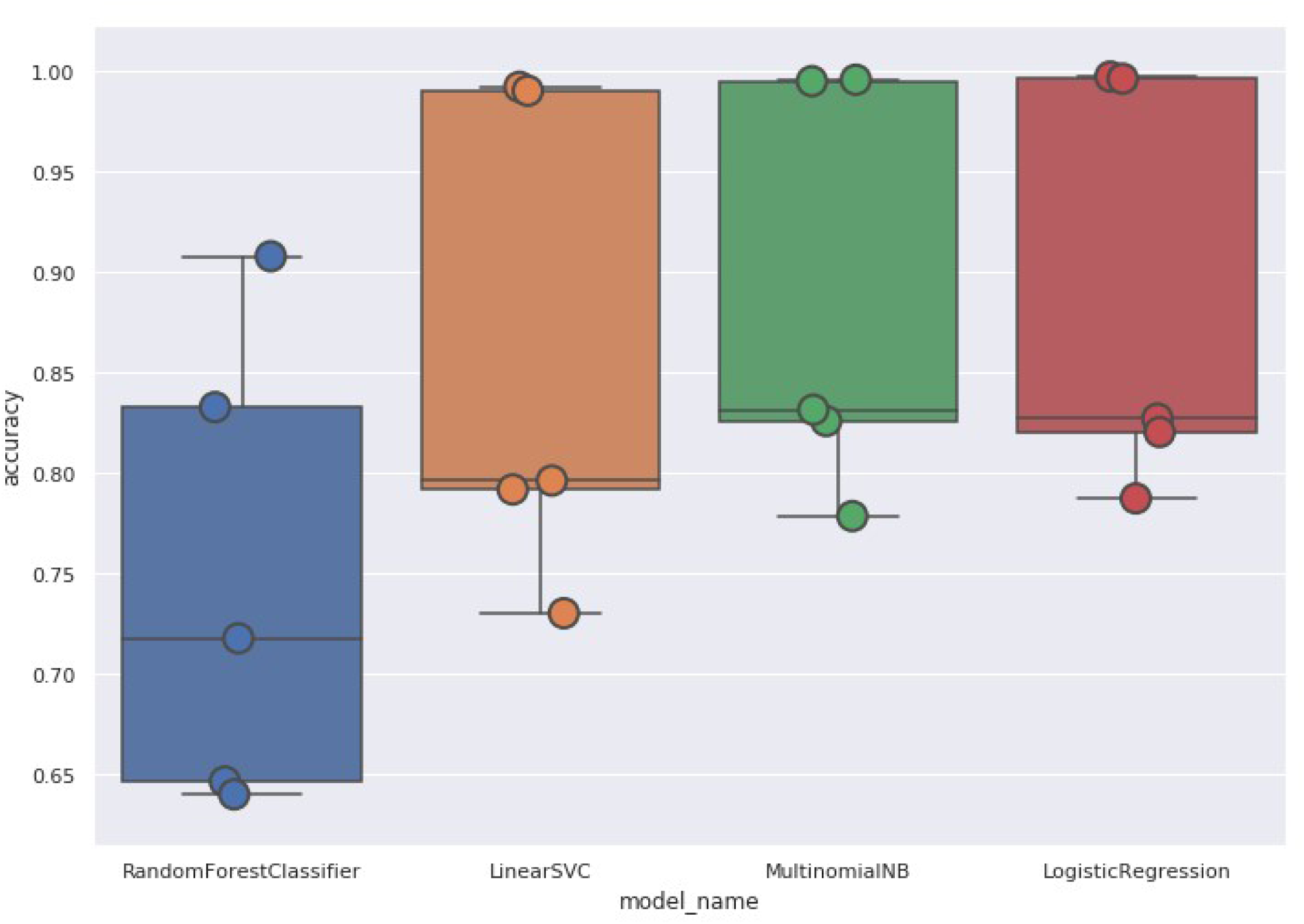
Box and whisker plot demonstrating the test-set classification accuracy (%; y-axis) for predicting the specific cochlear implant manufacturer after five-fold cross validation for each of the four machine algorithms. The lines within the boxes denote the mean classification accuracy.

**Figure 3.**
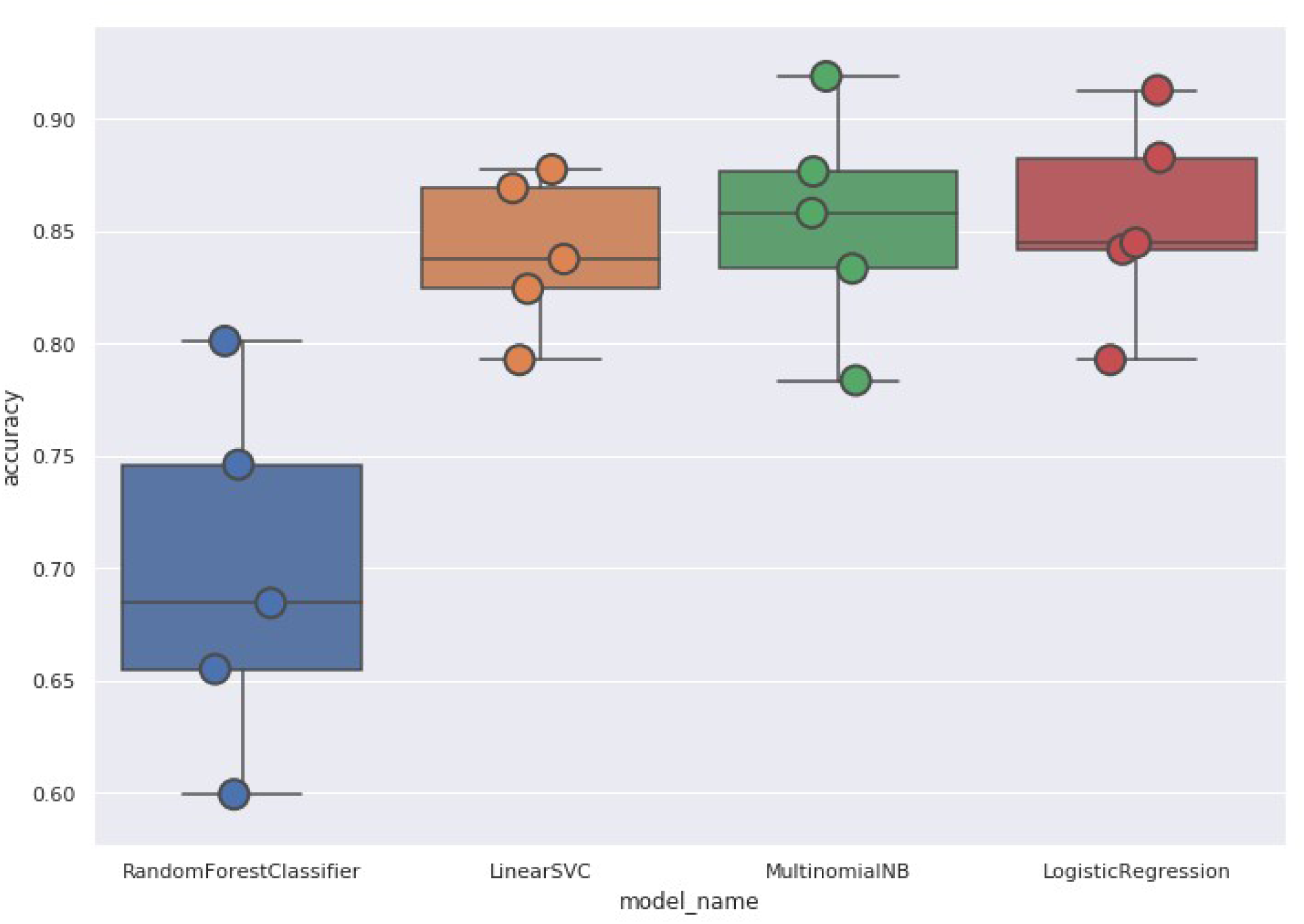
Box and whisker plot demonstrating the test-set classification accuracy (%; y-axis) for predicting adverse event type after five-fold cross validation for each of the four machine algorithms. The lines within the boxes denote the mean classification accuracy.

Computed classification metrics demonstrated the logistic regression model had the highest precision for MedEL and the highest recall for Cochlear Corporation (**Table 1**). In predicting the adverse event type, the linear SVC model achieved the highest precision for ‘injury’, whereas the logistic regression achieved the highest precision for ‘malfunction’. Several positive and negative predictors for each manufacturer and adverse event type were identified (**Table 2**). The logistic regression model was used to generate predictions on novel text examples for specific CI manufacturers (**Table 3**). The most influential positive predictors appear to represent the style in how the manufacturers structured their report. For instance, MDRs submitted by Advanced Bionics were predicted using words such as *“recipient’* and “pt” (assumed to be short form for ‘patient’). Whereas *“clinic”* and “*audiologist*’ were more predictive of Cochlear corporation MDRs. While these findings are not useful in identifying manufacturer-specific trends in adverse events, it is interesting that the algorithm identified an etymological style of reporting unique to each manufacturer.

**Table 1.**
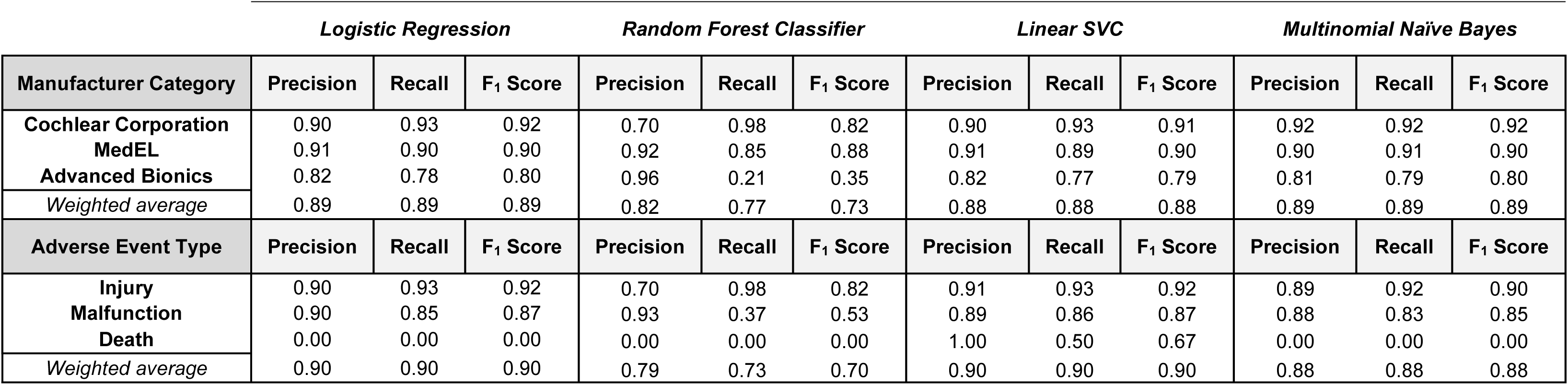
Machine learning model classification performance metrics. ***Weighted average**: averaging the support-weighted mean per label*.

|  | *Logistic Regression* |  |  | *Random Forest Classifier* |  |  | *Linear SVC* |  |  | *Multinomial Naïve Bayes* |  |  |
| --- | --- | --- | --- | --- | --- | --- | --- | --- | --- | --- | --- | --- |
| **Manufacturer Category** | **Precision** | **Recall** | **F<sub>1</sub> Score** | **Precision** | **Recall** | **F<sub>1</sub> Score** | **Precision** | **Recall** | **F<sub>1</sub> Score** | **Precision** | **Recall** | **F<sub>1</sub> Score** |
| **Cochlear Corporation** | 0.90 | 0.93 | 0.92 | 0.70 | 0.98 | 0.82 | 0.90 | 0.93 | 0.91 | 0.92 | 0.92 | 0.92 |
| **MedEL** | 0.91 | 0.90 | 0.90 | 0.92 | 0.85 | 0.88 | 0.91 | 0.89 | 0.90 | 0.90 | 0.91 | 0.90 |
| **Advanced Bionics** | 0.82 | 0.78 | 0.80 | 0.96 | 0.21 | 0.35 | 0.82 | 0.77 | 0.79 | 0.81 | 0.79 | 0.80 |
| *Weighted average* | 0.89 | 0.89 | 0.89 | 0.82 | 0.77 | 0.73 | 0.88 | 0.88 | 0.88 | 0.89 | 0.89 | 0.89 |
| **Adverse Event Type** | **Precision** | **Recall** | **F<sub>1</sub> Score** | **Precision** | **Recall** | **F<sub>1</sub> Score** | **Precision** | **Recall** | **F<sub>1</sub> Score** | **Precision** | **Recall** | **F<sub>1</sub> Score** |
| **Injury** | 0.90 | 0.93 | 0.92 | 0.70 | 0.98 | 0.82 | 0.91 | 0.93 | 0.92 | 0.89 | 0.92 | 0.90 |
| **Malfunction** | 0.90 | 0.85 | 0.87 | 0.93 | 0.37 | 0.53 | 0.89 | 0.86 | 0.87 | 0.88 | 0.83 | 0.85 |
| **Death** | 0.00 | 0.00 | 0.00 | 0.00 | 0.00 | 0.00 | 1.00 | 0.50 | 0.67 | 0.00 | 0.00 | 0.00 |
| *Weighted average* | 0.90 | 0.90 | 0.90 | 0.79 | 0.73 | 0.70 | 0.90 | 0.90 | 0.90 | 0.88 | 0.88 | 0.88 |

**Table 2.**
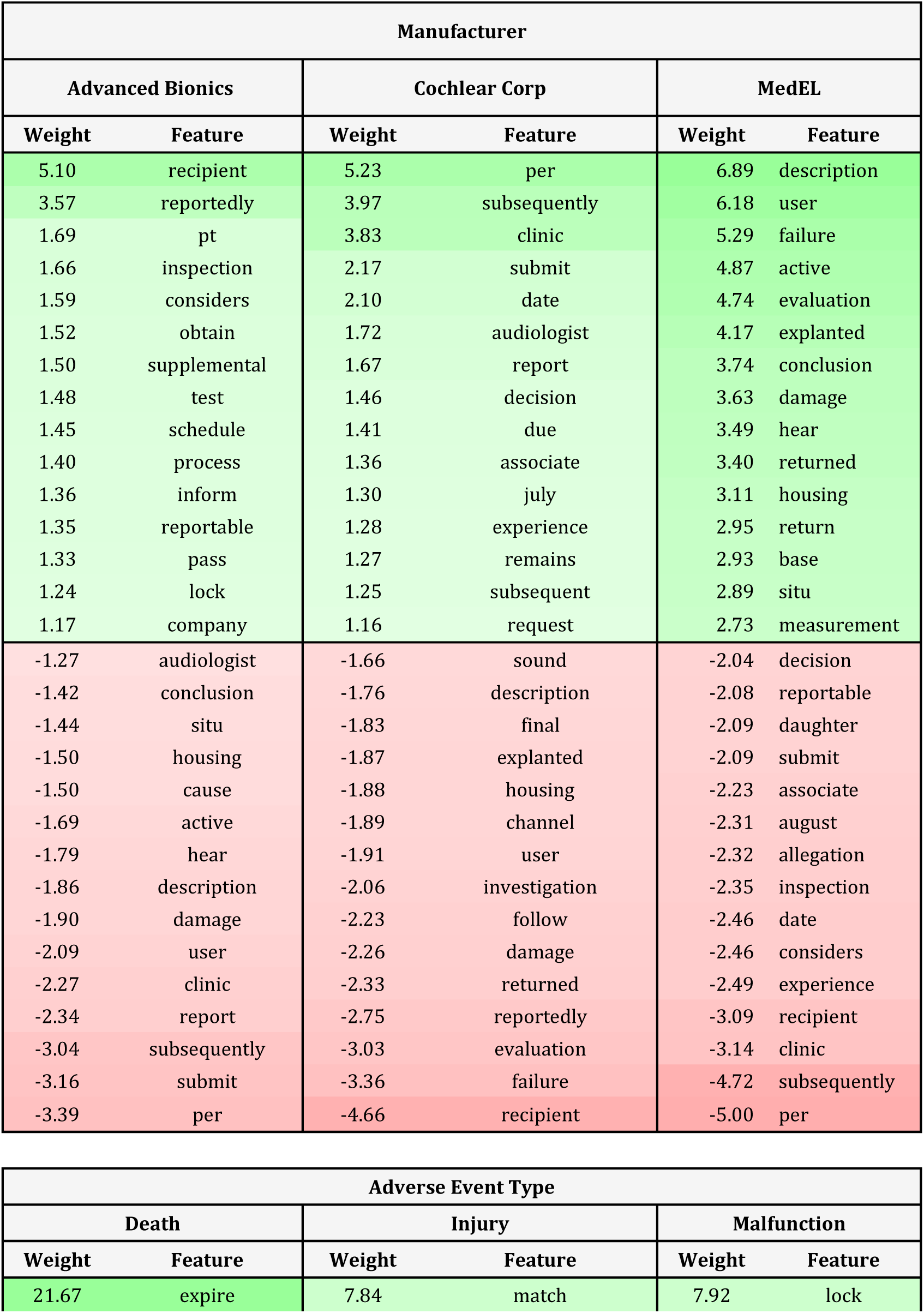
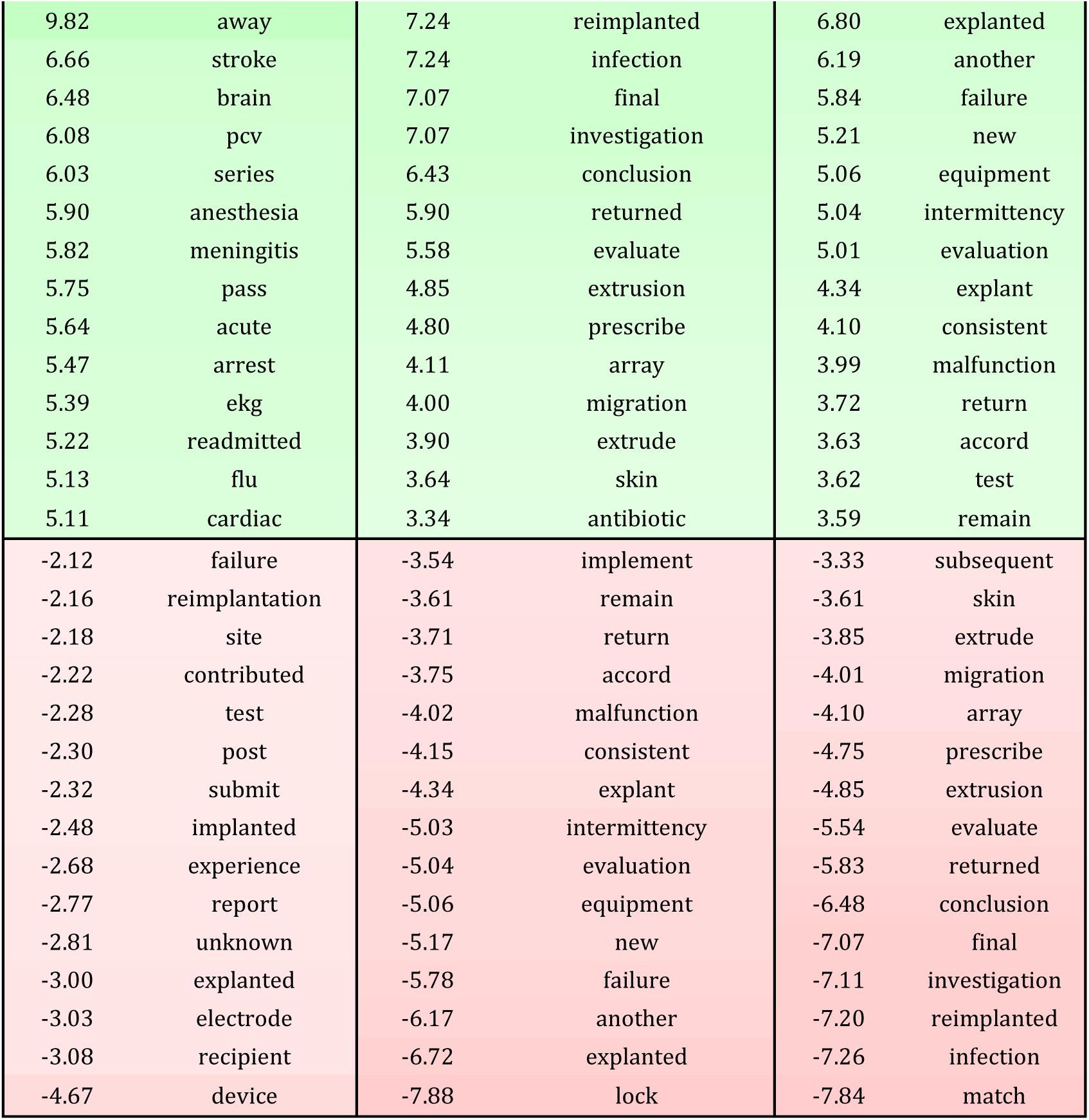
Top 15 most influential positive and negative predictive words for each manufacturer and adverse event type. Generated using the *ELI5* package.

| Manufacturer |  |  |  |  |  |
| --- | --- | --- | --- | --- | --- |
| Advanced Bionics |  | Cochlear Corp |  | MedEL |  |
| Weight | Feature | Weight | Feature | Weight | Feature |
| 5.10 | recipient | 5.23 | per | 6.89 | description |
| 3.57 | reportedly | 3.97 | subsequently | 6.18 | user |
| 1.69 | pt | 3.83 | clinic | 5.29 | failure |
| 1.66 | inspection | 2.17 | submit | 4.87 | active |
| 1.59 | considers | 2.10 | date | 4.74 | evaluation |
| 1.52 | obtain | 1.72 | audiologist | 4.17 | explanted |
| 1.50 | supplemental | 1.67 | report | 3.74 | conclusion |
| 1.48 | test | 1.46 | decision | 3.63 | damage |
| 1.45 | schedule | 1.41 | due | 3.49 | hear |
| 1.40 | process | 1.36 | associate | 3.40 | returned |
| 1.36 | inform | 1.30 | july | 3.11 | housing |
| 1.35 | reportable | 1.28 | experience | 2.95 | return |
| 1.33 | pass | 1.27 | remains | 2.93 | base |
| 1.24 | lock | 1.25 | subsequent | 2.89 | situ |
| 1.17 | company | 1.16 | request | 2.73 | measurement |
| -1.27 | audiologist | -1.66 | sound | -2.04 | decision |
| -1.42 | conclusion | -1.76 | description | -2.08 | reportable |
| -1.44 | situ | -1.83 | final | -2.09 | daughter |
| -1.50 | housing | -1.87 | explanted | -2.09 | submit |
| -1.50 | cause | -1.88 | housing | -2.23 | associate |
| -1.69 | active | -1.89 | channel | -2.31 | august |
| -1.79 | hear | -1.91 | user | -2.32 | allegation |
| -1.86 | description | -2.06 | investigation | -2.35 | inspection |
| -1.90 | damage | -2.23 | follow | -2.46 | date |
| -2.09 | user | -2.26 | damage | -2.46 | considers |
| -2.27 | clinic | -2.33 | returned | -2.49 | experience |
| -2.34 | report | -2.75 | reportedly | -3.09 | recipient |
| -3.04 | subsequently | -3.03 | evaluation | -3.14 | clinic |
| -3.16 | submit | -3.36 | failure | -4.72 | subsequently |
| -3.39 | per | -4.66 | recipient | -5.00 | per |

| Adverse Event Type |  |  |  |  |  |
| --- | --- | --- | --- | --- | --- |
| Death |  | Injury |  | Malfunction |  |
| Weight | Feature | Weight | Feature | Weight | Feature |
| 21.67 | expire | 7.84 | match | 7.92 | lock |
| 9.82 | away | 7.24 | reimplanted | 6.80 | explanted |
| 6.66 | stroke | 7.24 | infection | 6.19 | another |
| 6.48 | brain | 7.07 | final | 5.84 | failure |
| 6.08 | pcv | 7.07 | investigation | 5.21 | new |
| 6.03 | series | 6.43 | conclusion | 5.06 | equipment |
| 5.90 | anesthesia | 5.90 | returned | 5.04 | intermittency |
| 5.82 | meningitis | 5.58 | evaluate | 5.01 | evaluation |
| 5.75 | pass | 4.85 | extrusion | 4.34 | explant |
| 5.64 | acute | 4.80 | prescribe | 4.10 | consistent |
| 5.47 | arrest | 4.11 | array | 3.99 | malfunction |
| 5.39 | ekg | 4.00 | migration | 3.72 | return |
| 5.22 | readmitted | 3.90 | extrude | 3.63 | accord |
| 5.13 | flu | 3.64 | skin | 3.62 | test |
| 5.11 | cardiac | 3.34 | antibiotic | 3.59 | remain |
| -2.12 | failure | -3.54 | implement | -3.33 | subsequent |
| -2.16 | reimplantation | -3.61 | remain | -3.61 | skin |
| -2.18 | site | -3.71 | return | -3.85 | extrude |
| -2.22 | contributed | -3.75 | accord | -4.01 | migration |
| -2.28 | test | -4.02 | malfunction | -4.10 | array |
| -2.30 | post | -4.15 | consistent | -4.75 | prescribe |
| -2.32 | submit | -4.34 | explant | -4.85 | extrusion |
| -2.48 | implanted | -5.03 | intermittency | -5.54 | evaluate |
| -2.68 | experience | -5.04 | evaluation | -5.83 | returned |
| -2.77 | report | -5.06 | equipment | -6.48 | conclusion |
| -2.81 | unknown | -5.17 | new | -7.07 | final |
| -3.00 | explanted | -5.78 | failure | -7.11 | investigation |
| -3.03 | electrode | -6.17 | another | -7.20 | reimplanted |
| -3.08 | recipient | -6.72 | explanted | -7.26 | infection |
| -4.67 | device | -7.88 | lock | -7.84 | match |

**Table 3.**
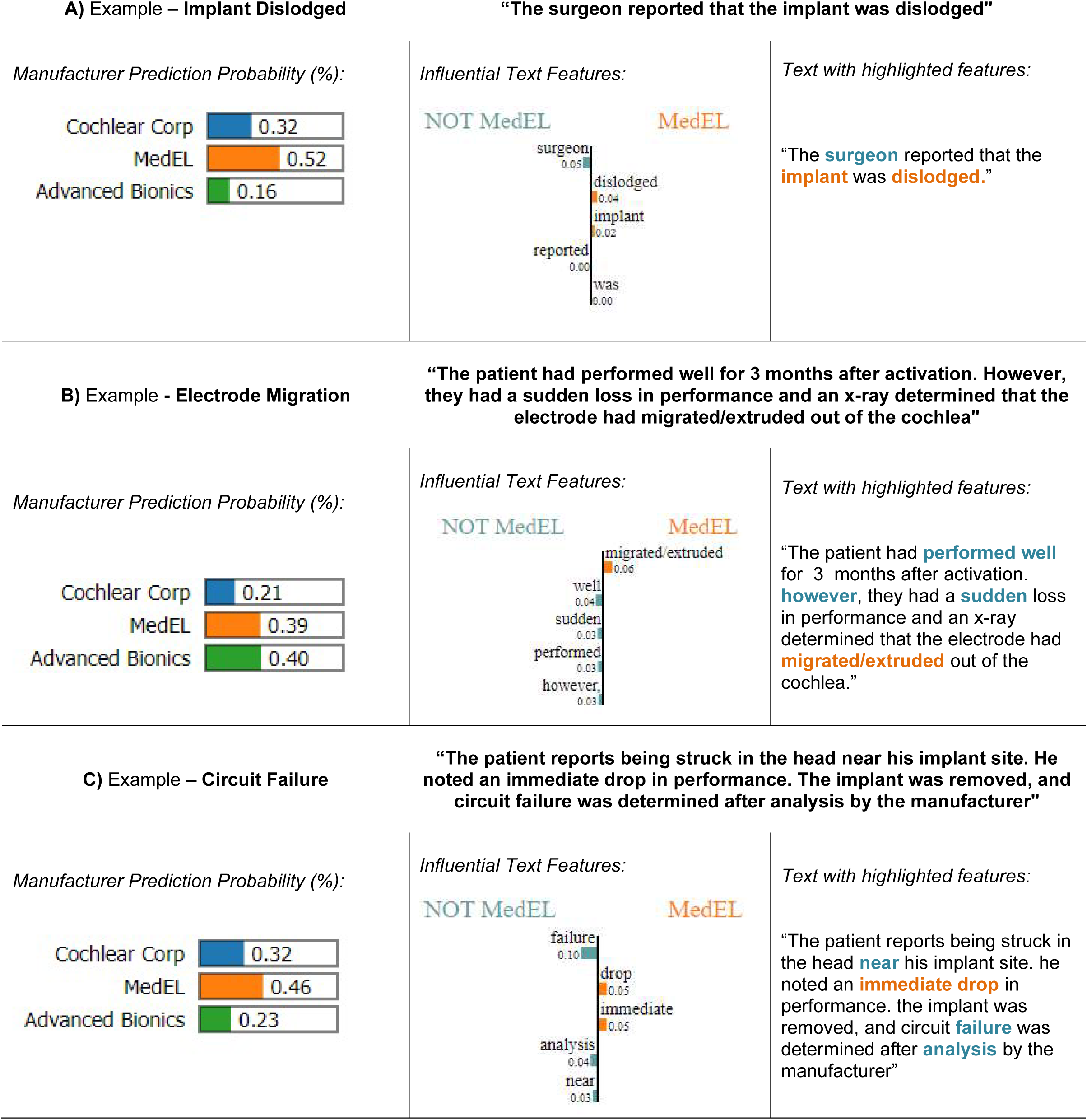
Manufacturer predictions using the trained logistic regression model on novel adverse event examples.

|  |  |  |
| --- | --- | --- |
| **A) Example – Implant Dislodged**<br>*Manufacturer Prediction Probability (%):*<br>Cochlear Corp 0.32<br>MedEL 0.52<br>Advanced Bionics 0.16 | **“The surgeon reported that the implant was dislodged”**<br>*Influential Text Features:* | *Text with highlighted features:*<br>“The **surgeon** reported that the **implant** was **dislodged**.” |
| **B) Example - Electrode Migration**<br>*Manufacturer Prediction Probability (%):*<br>Cochlear Corp 0.21<br>MedEL 0.39<br>Advanced Bionics 0.40 | **“The patient had performed well for 3 months after activation. However, they had a sudden loss in performance and an x-ray determined that the electrode had migrated/extruded out of the cochlea”**<br>*Influential Text Features:* | *Text with highlighted features:*<br>“The patient had **performed well** for 3 months after activation. **however**, they had a **sudden** loss in performance and an x-ray determined that the electrode had **migrated/extruded** out of the cochlea.” |
| **C) Example – Circuit Failure**<br>*Manufacturer Prediction Probability (%):*<br>Cochlear Corp 0.32<br>MedEL 0.46<br>Advanced Bionics 0.23 | **“The patient reports being struck in the head near his implant site. He noted an immediate drop in performance. The implant was removed, and circuit failure was determined after analysis by the manufacturer”**<br>*Influential Text Features:* | *Text with highlighted features:*<br>“The patient reports being struck in the head **near** his implant site. he noted an **immediate drop** in performance. the implant was removed, and circuit **failure** was determined after **analysis** by the manufacturer” |

## DISCUSSION

The objectives of this analysis were to apply natural language processing to the MAUDE MDR database to predict specific CI manufacturers and adverse event types based on the narrative text explanation of the adverse event. All the machine learning algorithms deployed against the MAUDE CI dataset achieved relatively high classification accuracy. While the model predictions themselves may hold little clinical utility, the insights derived from the model interpretations are useful in exploring the characteristics and quality of the narratives in the adverse reports. Interestingly, the most predictive words for each of the manufacturers resembled more of the writing style in the reporting adverse events, and less of patterns in specific clinically relevant adverse events. The narrative words predictive of specific adverse event types were intuitive and provided face validity support to our approach.

A recurring criticism of machine learning is that the algorithms are a ‘black box’ that lack practical interpretability. One of the benefits of using non-neural network algorithms, as we have chosen, is that analytic methods to interpret how the algorithm arrived at a specific prediction are available. We interpreted the logistic regression model’s predictions by tabulating the most predictive words calculated using a weighted measure of importance for each manufacturer and adverse event type. On our analysis of the most important narrative words, we believe the algorithm picked up on style of narrative writing rather than specific clinical concepts. Moreover, considering accuracy was high, we hypothesize that the manufacturers MDR narratives were internally consistent through the use of templates, report writing training, and/or the same personnel.

To further interrogate our algorithms’ ability to mine clinically useful trends in adverse event reports, we predicted the manufacturer base on our author-generated short adverse event narratives. In our examples, MedEL was predicted to be associated with the narratives describing CI dislodgement and circuit failure. For our example depicting electrode migration, the classification probability was approximately equal between MedEL and Advanced Bionics. Further analysis of these specific manufacturer adverse event relationships could be validated by cross-referencing specific model outcomes in independently published reports if they were to be made available. Because of the lack of standardization and weighting in reporting in the MAUDE database, we must strongly emphasize that we cannot make inferences or generalize these or other adverse events to a specific manufacturer.

Our study has important limitations that need mentioning. The most pressing limitation of our analysis is rooted in the circumstance that the MAUDE database is composed of non-standardized manufacturer-supplied adverse event reports. We cannot independently verify the accuracy of adverse events reports, nor assume that the adverse events are reported in a consistent manner. We assume that that the classification of the adverse event type is the appropriate label for each adverse event report. We cannot generalize on the frequency of specific adverse event types attributed to manufacturers and their devices. In particular, there are relatively few adverse event reports describing ‘death’ and thus our algorithms may fail to generalize to mortality reports when provided with new data. CI investigators have previously called for improvements to the adverse event reporting for CIs in the MAUDE database as well as guidelines for more specific detail in ‘who’ and ‘what’ should be contained within the reports.^45^ Anecdotal reports from manufacturers’ adverse events reporting compliance staff suggest that a standardized reporting system is under development with all three major CI manufacturers so that the adverse event report data will be more uniform in format and quality. We welcome this initiative that will enhance the quality and utility of the MAUDE database for future analyses.

Using supervised machine learning algorithms, our classification model was able to accurately predict the CI manufacturer based on adverse event text descriptions. From the interpretation of the model outputs, we believe the algorithms were able to accurately determine the adverse event based on the reporting style of the narrative. While we did not observe novel clinical concepts with specific association to manufacturers or adverse event types, we believe this shortcoming is less a function of our methodologic approach. Our attempt to derive clinically significant patterns in adverse events from this database was limited, in part, by the quality of the data inherent to the MAUDE database. Moreover, it is plausible that the pooled manufacturers adverse event data does not have sufficient granularity for which to tease out small differences between manufacturers. Compared to single institution cohort analyses, this ‘big data’ approach to an independent, national database may allow for further analysis of previously undiscovered or unreported trends in adverse events with specific manufacturers’ devices. Further work is underway to refine our approach to explore patterns at the device model level. We emphatically encourage efforts to standardize the CI database reporting methods to increase the quality of subsequent analyses.

## Data Availability

The data that support the findings of this study are openly available in the FDA MAUDE database at https://www.accessdata.fda.gov/scripts/cdrh/cfdocs/cfmaude/search.cfm.

## ACKNOWLEDGEMENTS

This work is supported in part by a 2018 Connaught Global Challenge Award from the University of Toronto.

